# A Semi-Automated Interactive Annotation Framework for Intraoperative Neurosurgical Ultrasound: A Proof-of-Concept Study

**DOI:** 10.64898/2026.10.03.26364663

**Authors:** Raphael Bertani, Siyar Bahadir, Marcio Yuri Ferreira, Leonardo Januário Campos Cardoso, Griffin Thomas, Netanel Ben-Shalom

## Abstract

**Objectives:** Intraoperative ultrasound (IOUS) is an increasingly valuable real-time imaging modality in neurosurgery, enabling visualization of brain parenchyma, tumor margins, ventricular anatomy, and vascular structures during cranial procedures. Despite its clinical relevance, the development of artificial intelligence (AI)-assisted interpretation tools for neurosurgical IOUS has been limited by the scarcity of large, annotated datasets. Manual pixel-level annotation of ultrasound video frames is labor-intensive, requires specialized neuroanatomical expertise, and scales poorly. To address this bottleneck, we developed a proof-of-concept interactive semi-automated annotation pipeline designed to explore the feasibility of scalable, expert-guided labeling for intraoperative neurosurgical IOUS.

**Methods:** We implemented the system as a napari-based graphical interface coupled with a scikit-learn Random Forest (RF) pixel classifier. Real intraoperative neurosurgical ultrasound videos from five craniotomy cases were used as training material. A neurosurgical expert annotated a minimal set of representative frames using brush-based labeling to define anatomical classes of interest. For each labeled pixel, a feature vector was computed comprising raw intensity, multi-scale Gaussian filtering (σ = 1, 2, 3), Laplacian of Gaussian, and Sobel edge magnitude. The RF classifier was trained on accumulated annotations using class-balanced weighting and out-of-bag (OOB) validation, then applied across full video sequences. An incremental accumulator module enabled multi-case training. Segmentation outputs were exported as TIFF overlays for expert quality review.

**Results:** Across five intraoperative cases, sparse expert annotation of a small number of representative frames per case was sufficient to train case-specific Random Forest classifiers that propagated labels across the full video sequences. Out-of-bag estimation served as an internal coherence check during training, and the classifier additionally produced per-class probability maps that exposed regions of uncertainty at acoustic boundaries. On expert qualitative review, the auto-segmented frames were anatomically coherent, with the classifier reliably distinguishing anechoic fluid compartments from echogenic tissue interfaces; residual misclassification was concentrated in low-contrast boundary regions.

**Conclusions:** This proof-of-concept study demonstrates that semi-automated, expert-guided annotation of intraoperative neurosurgical ultrasound is technically feasible using classical machine learning. By combining neurosurgical domain expertise with an interactive, incremental annotation workflow applied to real intraoperative acquisitions, this framework represents a foundational step toward scalable generation of labeled datasets for neurosurgical IOUS. Future directions include formal multi-rater validation, expanded multi-class anatomical labeling, and eventual migration to deep learning architectures as annotated datasets grow.

**Practitioner Points:**

- A napari-based interface coupled with a Random Forest pixel classifier can propagate sparse expert annotations across full intraoperative ultrasound video sequences, sharply reducing the manual labeling burden.
- The classifier’s per-class probability output highlights low-confidence, low-contrast acoustic boundaries, offering a built-in uncertainty signal that could inform real-time interpretation.
- This lightweight, GPU-free workflow provides a practical, scalable first step toward building annotated video datasets that could eventually support deep learning-based interpretation of neurosurgical IOUS.

## INTRODUCTION

Intraoperative ultrasound (IOUS) has emerged as an increasingly valuable imaging modality in neurosurgical practice. Its portability, cost-effectiveness, real-time acquisition capability, and seamless integration into the surgical workflow have made it a practical alternative or complement to intraoperative magnetic resonance imaging (iMRI) and neuronavigation systems [1–3]. In the context of brain tumor surgery, IOUS enables real-time visualization of tumor margins, residual tumor tissue, and surrounding anatomical structures, thereby informing intraoperative decision-making regarding the extent of resection [4,5]. Meta-analytic evidence suggests that IOUS-guided glioma resection is associated with improved gross total resection rates and overall survival compared with conventional neuronavigation alone [6,7].

Despite these clinical advantages, the widespread adoption and systematic exploitation of IOUS in neurosurgery remain constrained by several factors. IOUS image quality is highly variable and operator-dependent; acquisition angles, probe selection, and craniotomy constraints produce an effectively infinite space of possible brain views [8]. The resulting images are frequently degraded by artifacts, speckle noise, and unfamiliar perspectives, creating a steep learning curve that limits adoption even among experienced neurosurgeons [3,8]. These challenges are particularly pronounced in the context of non-navigated two-dimensional IOUS, which, despite being more accessible and cost-effective, demands greater interpretive expertise [9].

Artificial intelligence (AI) and machine learning (ML) hold considerable promise for mitigating these interpretability challenges. Deep learning-based segmentation models have demonstrated encouraging results in automatic glioma delineation from IOUS images, with multicenter studies reporting Dice coefficients above 0.70 for tumor segmentation [10–12]. However, the development of such models is fundamentally constrained by the availability of large, high-quality annotated datasets. The annotation bottleneck is widely recognized as a primary obstacle in medical image analysis: manual pixel-level labeling is labor-intensive, costly, and requires domain-specific expertise that cannot be easily crowd-sourced [13–15]. In the specific context of neurosurgical IOUS, this problem is compounded by the specialized neuroanatomical knowledge required to interpret ultrasound images, the heterogeneity of acquisition parameters across institutions, and the sheer volume of video data generated during each procedure.

Several strategies have been proposed to alleviate the annotation burden in medical imaging, including semi-supervised learning, active learning, transfer learning from pre-operative MRI annotations, and interactive annotation tools [14–17]. Platforms such as MONAI Label have demonstrated that AI-assisted interactive labeling can significantly reduce annotation times in three-dimensional medical imaging [18]. In the bioimage analysis community, tools like napari and ilastik have enabled accessible, interactive pixel classification using classical machine learning algorithms such as Random Forests, lowering the technical barrier for non-computational scientists [19,20]. To date, however, no interactive, semi-automated annotation pipeline has been specifically designed to address the unique challenges of intraoperative neurosurgical ultrasound.

In this study, we present a proof-of-concept interactive semi-automated annotation framework for intraoperative neurosurgical IOUS. The system integrates a napari-based graphical interface with a scikit-learn Random Forest pixel classifier, enabling a neurosurgical expert to annotate a minimal set of representative frames from real intraoperative ultrasound videos and propagate these annotations across full video sequences via machine-learning-assisted segmentation. The primary objective is not to propose a definitive segmentation solution, but to demonstrate the technical feasibility and practical viability of an incremental, expert-guided annotation workflow that could serve as a foundational building block for scalable labeled dataset generation in the neurosurgical IOUS domain.

## MATERIALS AND METHODS

### Ethics Statement

This retrospective proof-of-concept study used de-identified intraoperative ultrasound recordings obtained during routine clinical care, with no additional patient procedures or changes to clinical management. Institutional review board or ethics committee review was not sought.

### Data Source and Case Selection

Intraoperative neurosurgical ultrasound videos were sourced from intraoperative acquisitions performed at the participating institution. Five consecutive cases involving intraoperative B-mode ultrasound were included in this proof-of-concept study. Cases were selected to represent a range of pathologies and anatomical contexts encountered during routine neurosurgical practice. Ultrasound acquisitions were performed with a standard intraoperative B-mode system, with imaging obtained through the craniotomy window. Video sequences were exported as individual frames for processing.

### System Architecture

The annotation framework was implemented as a Python-based pipeline integrating three principal components: (1) an interactive visualization and annotation interface built on the napari multi-dimensional image viewer; (2) a pixel-level feature extraction module; and (3) a Random Forest (RF) classifier from the scikit-learn library [21,22]. The system was designed to operate on standard hardware without requiring GPU acceleration, emphasizing accessibility for clinical research settings. An overview of the full pipeline is shown in Figure 1.

**Figure 1.**
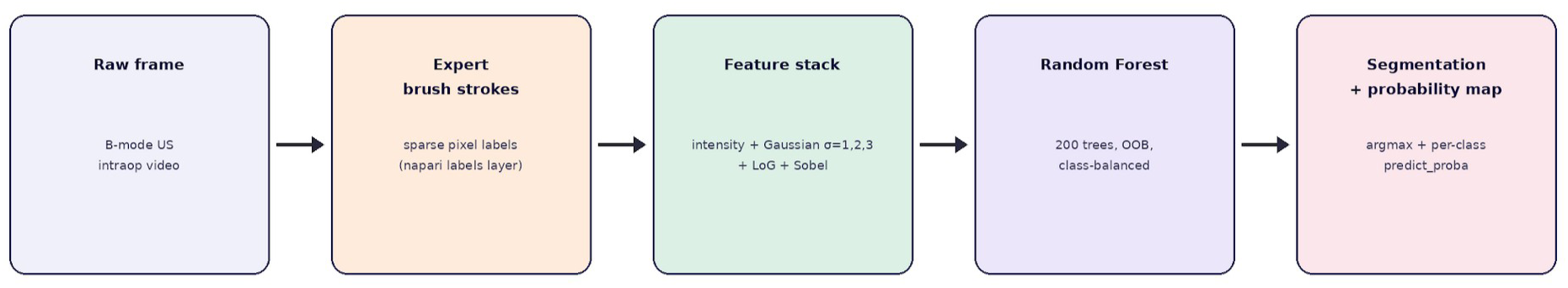
Overview of the semi-automated annotation pipeline. Each B-mode video frame passes through five stages: (1) raw frame loaded into napari; (2) sparse expert brush-stroke annotation; (3) multi-scale feature extraction (raw intensity, Gaussian σ = 1, 2, 3, Laplacian of Gaussian, Sobel); (4) Random Forest training with out-of-bag (OOB) validation and class-balanced weighting; (5) prediction across the full video sequence, generating both hard segmentation masks and per-class probability maps.

### Interactive Annotation Workflow

For each case, the ultrasound video was loaded into the napari viewer as a multi-frame image stack. The neurosurgical expert (R.B., a board-certified neurosurgeon experienced in IOUS interpretation) reviewed the video sequence and selected a minimal number of representative frames for manual annotation. Using the napari brush-based labeling tool, the expert painted regions of interest directly onto a labels layer, assigning each pixel to one of the predefined anatomical classes. The classes of interest were defined based on the anatomical structures visible in the specific case and included, but were not limited to: tumor tissue, normal brain parenchyma, ventricular walls, echogenic interfaces, falx cerebri, and background. The annotation strategy prioritized sparse but representative labeling: rather than exhaustively segmenting entire frames, the expert annotated small, diagnostically informative regions that captured each class’s acoustic signature and textural characteristics.

### Feature Extraction

For each labeled pixel, a feature vector was computed to capture local intensity and textural information at multiple spatial scales. The feature set comprised: (1) raw pixel intensity; (2) Gaussian-filtered intensity at three scales (σ = 1, 2, and 3 pixels), capturing local brightness at progressively coarser resolutions; (3) Laplacian of Gaussian (LoG) response, encoding second-order intensity variations sensitive to edges and boundaries; and (4) Sobel edge magnitude, providing gradient-based edge detection. This six-dimensional feature vector was designed to capture the multi-scale intensity and textural signatures characteristic of different tissue types in neurosurgical ultrasound. Features were computed using standard implementations from the scikit-image library [23].

### Classifier Training and Prediction

The RF classifier was configured with an ensemble of decision trees and trained using class-balanced sample weighting to mitigate the effect of imbalanced class representation in the sparse annotations. Out-of-bag (OOB) estimation was enabled to provide an internal cross-validation metric without requiring a held-out validation set, which is particularly valuable given the limited number of annotated frames. The classifier was trained on the concatenated feature vectors from all labeled pixels across the annotated frames for a given case. Upon completion of training, the model was applied to predict pixel-wise class assignments for every frame in the video sequence, generating a full segmentation overlay for the entire case. In addition to the argmax hard segmentation, the trained classifier outputs a per-class probability for every pixel via predict_proba. These probability maps were retained for qualitative inspection of model confidence and to visualize regions of class ambiguity at acoustic boundaries (Figure 3).

### Incremental Multi-Case Training

A key design feature of the framework is an incremental accumulator module that enables progressive model refinement across multiple cases. When processing a new case, the expert’s annotations are appended to the accumulated feature-label pairs from previously processed cases. The RF classifier is then retrained on the expanded dataset, progressively broadening its exposure to the anatomical, textural, and acoustic variability inherent in neurosurgical IOUS across different patients, pathologies, probe orientations, and ultrasound systems. Trained models are serialized using Python’s joblib library for batch deployment across the dataset.

### Output and Quality Review

Segmentation outputs were exported as multi-frame TIFF overlay stacks, with each pixel assigned a class-specific color map for visual inspection. The neurosurgical expert reviewed all auto-segmented frames within the napari viewer, assessing the anatomical coherence of the predicted segmentations against the underlying ultrasound images. This qualitative review served as the primary quality metric in this proof-of-concept study. No formal quantitative validation against independently generated ground-truth masks was performed, as this was explicitly outside the scope of the current feasibility study.

## RESULTS

### Case Characteristics

Five intraoperative B-mode ultrasound cases were processed through the pipeline. The cases spanned a range of pathologies and anatomical contexts: three were intracranial tumor acquisitions of varying internal architecture, and two represented ventricular and catheter-trajectory views. This deliberate heterogeneity exposed the classifier to a spectrum of acoustic appearances, from multi-cystic lesions with hyperechoic septations to predominantly anechoic ventricular compartments.

### Annotation Efficiency

For each case, only a small number of representative frames required manual expert annotation, and within those frames the expert labeled sparse, diagnostically informative regions rather than exhaustively segmenting every pixel. This sparse-labeling strategy meant that the expert effort needed to seed a case-specific classifier was a small fraction of that demanded by exhaustive, frame-by-frame manual segmentation of the full video sequences. Once trained, the classifier propagated these sparse annotations across every frame of the corresponding case without further expert input, so the marginal cost of labeling additional frames within a case approached zero.

### Classifier Behavior

Out-of-bag estimation, enabled during training, provided an internal coherence signal that tracked the separability of the annotated classes without requiring a held-out validation set. Beyond the hard (argmax) segmentation, the classifier produced per-class probability maps via predict_proba; these maps localized classifier confidence and made regions of class ambiguity at acoustic boundaries directly visible (Figure 3). In practice, the probability output was highest within the interior of well-defined anechoic and hyperechoic regions and lowest along the transitional interfaces between them.

### Qualitative Segmentation Assessment

On expert review, the auto-segmented frames were anatomically coherent across the five cases. The classifier consistently distinguished anechoic fluid compartments from echogenic tissue interfaces, recovering the principal structures visible in each acquisition. A representative tumor case is shown in Figure 2, in which the classifier recovered the multiple anechoic compartments, the hyperechoic septations and walls, and a surrounding cortical band of a multi-cystic lesion. The probabilistic output for one of the tumor cases is illustrated in Figure 3, and frame-level segmentation across all five cases is shown in Figure 4. For Case 5 (VT2), segmentation coherence visibly improved once the case was incorporated into the incremental accumulator, with sharper recovery of the catheter trajectory and the surrounding fluid compartment relative to the earlier single-case output (Figure 5). Residual misclassification was concentrated in low-contrast boundary regions, near-field artifact, and acoustic shadow zones, where class signatures overlap and the probability maps correspondingly indicated lower confidence.

**Figure 2.**
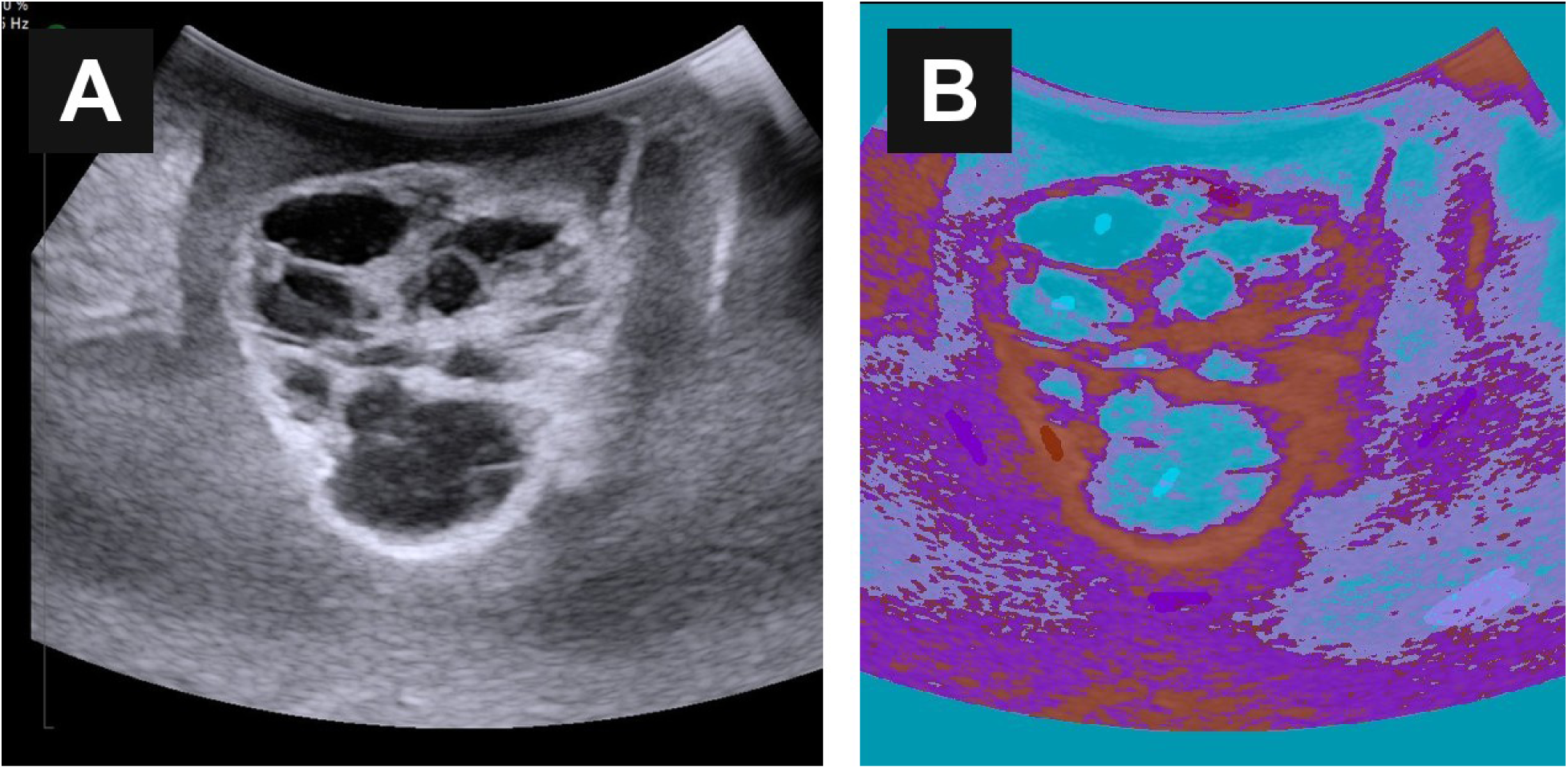
Representative case (Case 3), a patient with a multi-cystic intracranial lesion. A, Raw B-mode sonogram demonstrating the lesion. B, Random Forest segmentation output showing recovery of the multiple anechoic compartments (cyan, fluid class), the hyperechoic septations and walls (burgundy, tumor/hyperechoic-tissue class), and a surrounding cortical band (violet). Class color code: burgundy = tumor/hyperechoic tissue; cyan = fluid; violet = cortex; purple = edema.

**Figure 3.**
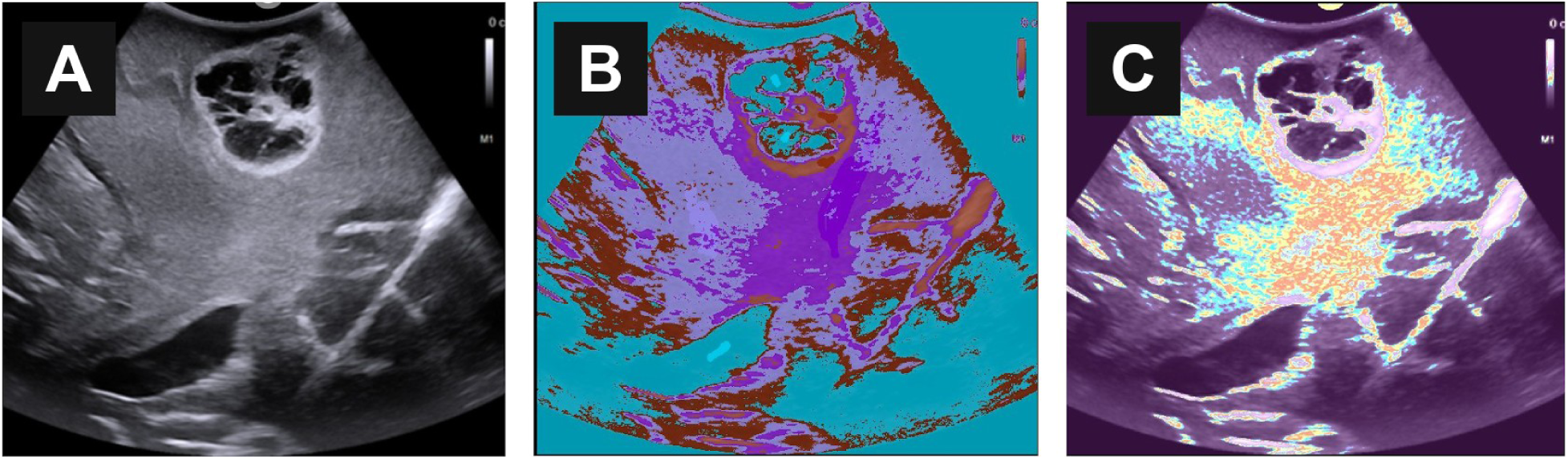
Case 2, a patient undergoing intraoperative ultrasound-guided tumor resection — raw frame, hard segmentation, and per-class probability map. A, Raw B-mode sonogram. B, Hard (argmax) segmentation output. C, Per-class probability map derived from predict_proba; warmer colors indicate higher classifier confidence and cooler colors mark regions of class-boundary uncertainty. The probability output provides a natural visualization of regions where the classifier is uncertain, typically at acoustic interfaces.

**Figure 4.**
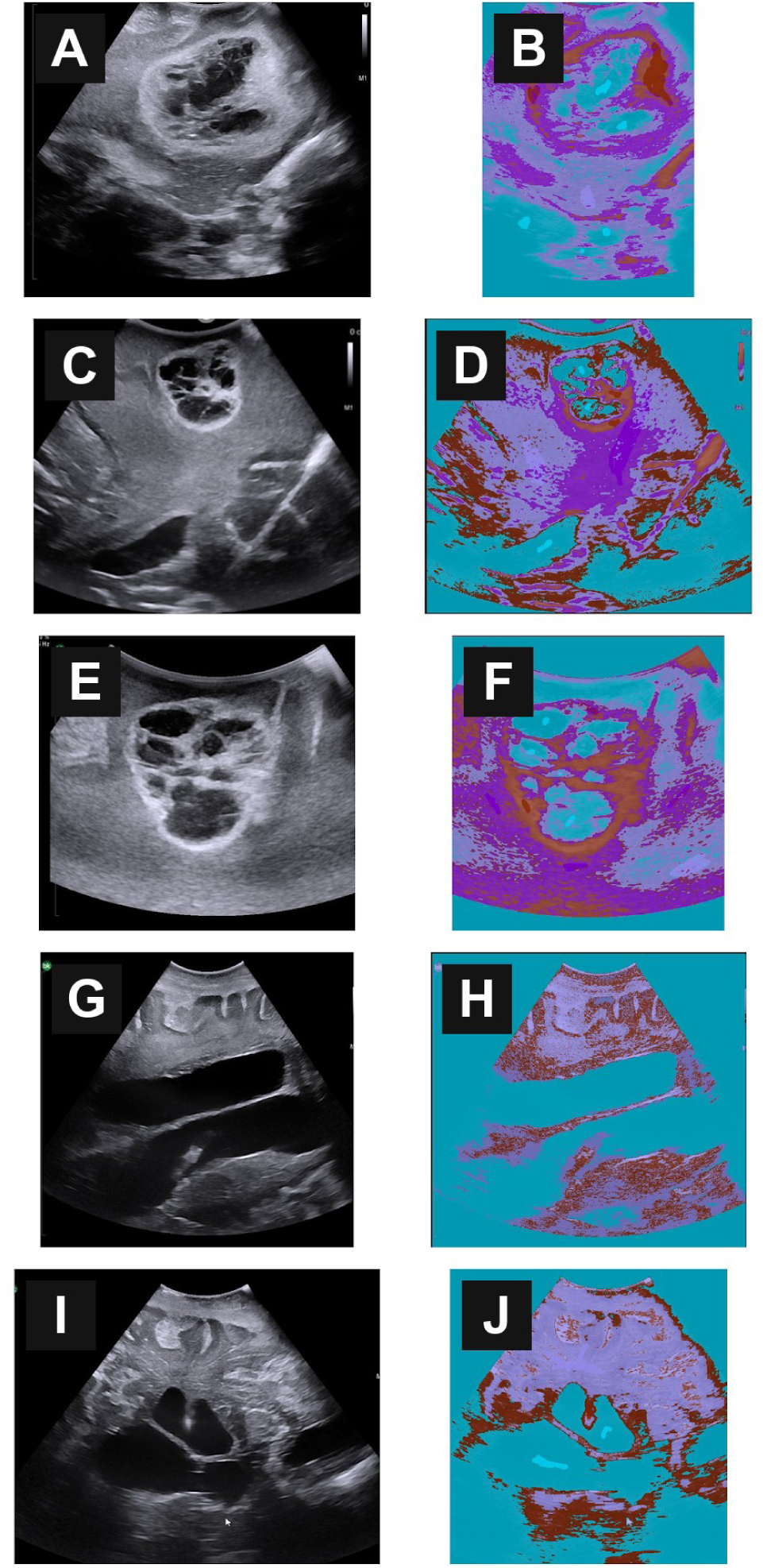
Frame-level segmentation outputs across the five included cases. A and B, Case 1. C and D, Case 2. E and F, Case 3. G and H, Case 4. I and J, Case 5. For each case, the first (odd-lettered) panel shows the raw B-mode frame and the second (even-lettered) panel shows the corresponding Random Forest segmentation output. Cases 1, 2, and 3 represent intracranial tumor acquisitions of varying internal architecture; Cases 4 and 5 represent ventricular and catheter-trajectory views. Across all five cases, the classifier consistently identifies anechoic fluid compartments (cyan) and echogenic tissue interfaces (burgundy), with characteristic class-boundary noise that is most prominent in regions of low acoustic contrast.

**Figure 5.**
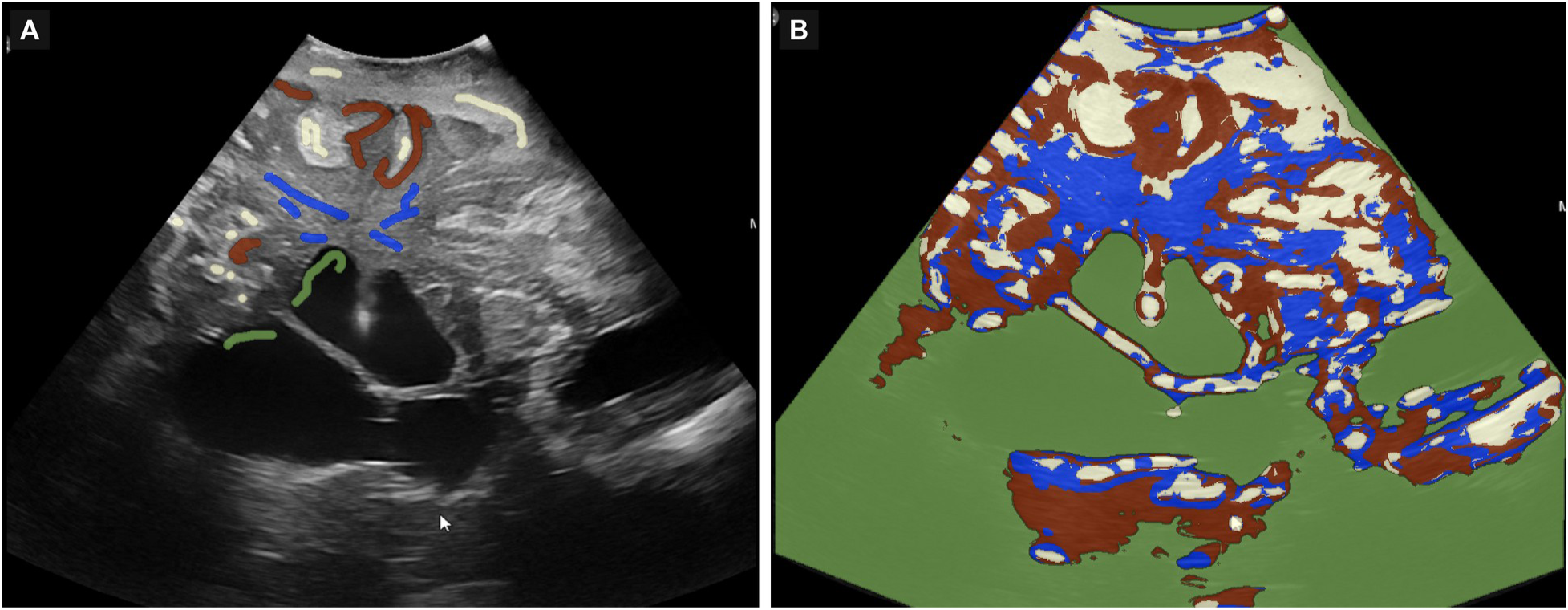
Case 5 (VT2), a patient with a ventricular catheter, improved segmentation output. A, Raw B-mode sonogram with sparse expert annotations. B, Random Forest segmentation after this case was incorporated into the incremental accumulator, showing improved separation of acoustic tissue classes relative to the earlier single-case output. Class color code: brown and blue = white matter; green = fluid; white = gray matter. The catheter was not assigned a separate class and was clearly visible as a protrusion into the ventricle.

## DISCUSSION

This study demonstrates the technical feasibility and practical viability of a semi-automated, expert-guided annotation workflow for intraoperative neurosurgical ultrasound. Using a lightweight classical machine learning approach, a Random Forest pixel classifier operating on handcrafted multi-scale intensity and texture features, the framework produced anatomically coherent segmentations when guided by sparse annotations from a neurosurgical domain expert. The deliberate use of classical ML rather than deep learning was a design choice appropriate for this stage of development: Random Forest classifiers are interpretable, require no GPU infrastructure, train in seconds, and perform acceptably well on small annotated datasets where deep learning architectures would overfit [19,20]. This restraint is consistent with recent critical appraisals cautioning that the clinical evidence for AI and deep-learning tools in intraoperative neurosurgery still lags the surrounding enthusiasm [27].

The annotation bottleneck represents a fundamental challenge in medical image analysis and is particularly acute in neurosurgical IOUS. Unlike other imaging domains in which crowdsourced annotations are feasible, IOUS interpretation requires specialized neuroanatomical expertise, knowledge of the acoustic properties of brain tissue, and familiarity with the specific artifact signatures of intraoperative ultrasound [3,8,13]. The framework presented here directly addresses this bottleneck by minimizing the number of expert-annotated frames required while maximizing the propagation of domain knowledge through ML-assisted segmentation. This approach is philosophically aligned with active learning and human-in-the-loop annotation paradigms that have demonstrated efficiency gains in medical image labeling [14,18].

The multicenter datasets that currently exist for neurosurgical IOUS — notably BraTioUS, comprising 1,669 images from 142 patients across six centers, and the RESECT database of pre-operative MRI and intraoperative ultrasound in low-grade gliomas — represent important milestones in the field [10,24,25]. However, these datasets primarily focus on glioma segmentation and provide static 2D images rather than continuous video sequences captured during real-world surgical procedures. Our framework is designed to operate on video-level data, generating frame-by-frame segmentation across entire surgical sequences, thereby complementing existing datasets by providing temporally resolved annotations.

The probabilistic nature of the Random Forest output is itself an asset: pixel-wise class probabilities provide a natural uncertainty signal at acoustic boundaries (Figure 3), which a future implementation could surface to the operator as a confidence overlay during real-time interpretation. This probabilistic transparency is a methodological advantage over hard-segmentation deep learning models that do not natively expose calibrated per-pixel confidence scores without additional post hoc calibration. The catheter-trajectory case (VT2) is particularly relevant in this regard, as prior work has shown that deep-learning approaches can detect foreign-body objects and resolve discrete neural structures on intraoperative ultrasound in real time [26,28]; a probabilistic pixel classifier offers a lightweight, interpretable complement to those models for highlighting catheter and instrument boundaries.

Several important limitations of this proof-of-concept study must be acknowledged. First, no formal quantitative validation against independently generated ground-truth masks was performed; the quality assessment was limited to expert qualitative review. Second, the study included only five intraoperative cases from a single institution, which is insufficient to assess the generalizability of the trained models across different ultrasound systems, probe configurations, and surgical contexts. Third, no matching post-operative imaging was available for these cases, precluding correlation between the IOUS segmentation outputs and post-operative MRI-based tumor delineation. Fourth, the feature set employed was deliberately simple and may not capture the full range of textural and contextual information available in neurosurgical ultrasound. Fifth, inter-rater reliability was not assessed, as a single expert performed all annotations.

However, the current work explicitly validates the concept of an interactive, incremental annotation workflow rather than proposing a definitive segmentation solution. As the annotated dataset grows through continued application of this pipeline, several natural extensions become feasible: formal multi-rater validation studies to assess annotation consistency; expansion of the multi-class anatomical labeling scheme; integration of direct database ingestion pipelines; and, critically, migration from classical pixel classifiers to deep learning architectures, such as U-Net variants, which have demonstrated strong performance in IOUS tumor segmentation [11,12] as the volume of labeled data reaches the threshold where such architectures can be effectively trained.

## CONCLUSION

This proof-of-concept study demonstrates that semi-automated, expert-guided annotation of intraoperative neurosurgical ultrasound is technically feasible and practically viable using classical machine learning. By combining neurosurgical domain knowledge with an interactive Random Forest pixel-classification workflow applied to real intraoperative acquisitions, the framework takes a foundational step toward scalable labeled dataset generation for neurosurgical IOUS. The system is deliberately lightweight, interpretable, and accessible — requiring no GPU infrastructure and operating on standard clinical research hardware. While the current work is limited in scope and explicitly validates the annotation workflow rather than proposing a definitive segmentation tool, we believe this pipeline represents a necessary first building block toward higher-accuracy intraoperative ultrasound interpretation. As annotated datasets grow through the incremental application of this framework, migration to deep learning architectures capable of autonomously interpreting IOUS may become possible, ultimately contributing to improved intraoperative decision-making and surgical outcomes.

## Data Availability

All data produced in the present study are available upon reasonable request to the authors

## ACKNOWLEDGMENTS

The authors have no additional acknowledgments to disclose beyond the contributions of the listed authors. This work has not been previously presented at any meeting or conference.

## Funding

The authors declare that no funds, grants, or other support were received during the preparation of this manuscript.

## Disclosures

The authors declare no conflicts of interest to disclose.

## Artificial Intelligence Generated Content

No generative AI tools were used to draft, generate, or analyze the scientific content, data, or images of this manuscript. Language-editing assistance, if any, was limited to grammar and spelling checks.

## Notes

### Competing Interest Statement

The authors have declared no competing interest.

## REFERENCES

1. Dixon L, Lim A, Grech-Sollars M, Nandi D, Camp SJ. Intraoperative ultrasound in brain tumor surgery: a review and implementation guide. Neurosurg Rev. 2022;45(4):2503–2515.

2. Moiyadi AV. Intraoperative ultrasound technology in neuro-oncology practice — current role and future applications. World Neurosurg. 2016;93:81–93.

3. Sastry R, Bi WL, Pieper S, et al. Applications of ultrasound in the resection of brain tumors. J Neuroimaging. 2017;27(1):5–15.

4. Wu H, Cheng Y, Gao W, et al. Progress in the application of ultrasound in glioma surgery. Front Med (Lausanne). 2024;11:1388728.

5. Cepeda S, García-García S, Arrese I, Sarabia R. Non-navigated 2D intraoperative ultrasound: an unsophisticated surgical tool to achieve high standards of care in glioma surgery. J Neurooncol. 2024;167(3):387–396.

6. Pichardo-Rojas PS, Zarate C, Arguelles-Hernández J, et al. Intraoperative ultrasound for surgical resection of high-grade glioma and glioblastoma: a meta-analysis of 732 patients. Neurosurg Rev. 2024;47(1):120.

7. Mahboob S, McPhillips R, Qiu Z, et al. Intraoperative ultrasound-guided resection of gliomas: a meta-analysis and review of the literature. World Neurosurg. 2016;92:255–263.

8. Pino MA, Imperato A, Musca I, et al. New hope in brain glioma surgery: the role of intraoperative ultrasound. A review. Brain Sci. 2018;8(11):202.

9. West TR, Mazurek MH, Perez NA, et al. Navigated intraoperative ultrasound offers effective and efficient real-time analysis of intracranial tumor resection and brain shift. Oper Neurosurg. 2025;28(2):148–158.

10. Cepeda S, García-García S, Arrese I, et al. Deep learning-based glioma segmentation of 2D intraoperative ultrasound images: a multicenter study using the Brain Tumor Intraoperative Ultrasound Database (BraTioUS). Cancers. 2025;17(2):315.

11. Carton FX, Chabanas M, Munkvold BKR, Reinertsen I, Noble JH. Automatic segmentation of brain tumor resections in intraoperative ultrasound images using U-Net. J Med Imaging (Bellingham). 2020;7(3):031503.

12. Weld A, Agrawal A, Giannarou S. Ultrasound segmentation using a 2D UNet with Bayesian volumetric support. In: Xiao Y, Yang G, Song S, eds. Lesion Segmentation in Surgical and Diagnostic Applications. Lecture Notes in Computer Science. Vol 13648. Springer; 2023.

13. Singh P, Cirrone J, Solares R, et al. Shifting to machine supervision: annotation-efficient semi and self-supervised learning for automatic medical image segmentation and classification. Sci Rep. 2024;14(1):11091.

14. Yu L, Chen X. Label-efficient deep learning in medical image analysis: challenges and future directions. arXiv preprint arXiv:2303.12484. 2025.

15. Diaz-Pinto A, Alle S, Nath V, et al. MONAI Label: a framework for AI-assisted interactive labeling of 3D medical images. Med Image Anal. 2024;95:103207.

16. Tajbakhsh N, Jeyaseelan L, Li Q, Chiang JN, Wu Z, Ding X. Embracing imperfect datasets: a review of deep learning solutions for medical image segmentation. Med Image Anal. 2020;63:101693.

17. Haan Münch J, Arntzen K, Bjørnerud A, et al. Automatic brain tumor segmentation in 2D intra-operative ultrasound images using MRI tumor annotations. Cancers. 2025;17(11):1840.

18. Diaz-Pinto A, Alle S, Nath V, et al. MONAI Label: a framework for AI-assisted interactive labeling of 3D medical images. arXiv preprint arXiv:2203.12362. 2022.

19. Haase R, Royer LA, Steinbach P, et al. CLIJ: GPU-accelerated image processing for everyone. Nat Methods. 2020;17(1):5–6.

20. Berg S, Kutra D, Kroeger T, et al. ilastik: interactive machine learning for (bio)image analysis. Nat Methods. 2019;16(12):1226–1232.

21. Pedregosa F, Varoquaux G, Gramfort A, et al. Scikit-learn: machine learning in Python. J Mach Learn Res. 2011;12:2825–2830.

22. Sofroniew N, Lambert T, Evans K, et al. napari: a multi-dimensional image viewer for Python. Zenodo. 2022. doi:10.5281/zenodo.3555620.

23. van der Walt S, Schönberger JL, Nunez-Iglesias J, et al. scikit-image: image processing in Python. PeerJ. 2014;2:e453.

24. Cepeda S, García-García S, Arrese I, et al. BraTioUS: a multicenter dataset of baseline intraoperative brain tumor ultrasound images. Data Brief. 2026;60:111364.

25. Xiao Y, Fortin M, Unsgård G, Rivaz H, Reinertsen I. Retrospective evaluation of cerebral tumors (RESECT): a clinical database of pre-operative MRI and intra-operative ultrasound in low-grade glioma surgeries. Med Phys. 2017;44(7):3875–3882.

26. Foley KT, Mroz TE, Arnold PM, et al. Artificial intelligence-enabled, real-time intraoperative ultrasound imaging of neural structures within the psoas: validation in a porcine spine model. Spine (Phila Pa 1976). 2021;46(3):E146–E152.

27. Patel T, et al. Critical appraisal of artificial intelligence and deep-learning tools for intraoperative neurosurgery: hype versus evidence. Int J Surg. 2026;112(2):e1234.

28. Abramson HG, Curry EJ, Mess G, et al. Automatic detection of foreign body objects in neurosurgery using a deep learning approach on intraoperative ultrasound images: from animal models to first in-human testing. Front Surg. 2022;9:1040066.

